# Multivariate Echocardiographic Phenotyping of Hypertensive Heart Failure Using Unsupervised Machine Learning: A Pilot Study

**DOI:** 10.64898/2026.06.20.26356124

**Authors:** Joseph Onyemachi, James Chiedozie Maduka

## Abstract

**Background:** Heart failure in hypertensive patients is heterogeneous and poorly captured by traditional left ventricular ejection fraction (LVEF)-based classification. Multivariate echocardiographic data combined with unsupervised machine learning may provide a more precise phenotypic characterization. This pilot study evaluated the feasibility of unsupervised clustering of routine transthoracic echocardiographic data to identify phenotypic subgroups of hypertensive heart failure.

**Methods:** This retrospective pilot study analyzed transthoracic echocardiography reports from hypertensive patients with clinical heart failure. After data cleaning and exclusion of incomplete records, 102 patients with 11 echocardiographic variables were included. Variables describing left ventricular geometry, systolic function, and diastolic performance were standardized and subjected to K-means clustering. Optimal cluster number was determined using the elbow method and silhouette analysis. Cluster characteristics were assessed using descriptive statistics and Kruskal–Wallis testing. Concordance with LVEF-based heart failure categories was evaluated.

**Results:** Three distinct echocardiographic phenotypes were identified. Cluster 0 (n = 50) demonstrated preserved LVEF with concentric remodeling, consistent with heart failure with preserved ejection fraction (HFpEF) phenotype. Cluster 1 (n = 37) showed marked ventricular dilation and reduced systolic function, consistent with heart failure with reduced ejection fraction (HFrEF). Cluster 2 (n = 15) exhibited concentric hypertrophy with intermediate LVEF, consistent with heart failure with mildly reduced ejection fraction (HFmrEF)-like phenotype. All echocardiographic variables differed significantly across clusters (p < 0.001). While Cluster 0 showed strong concordance with HFpEF (96%), Clusters 1 and 2 demonstrated substantial overlap across LVEF categories, indicating partial discordance between structural phenotypes and LVEF-based classification.

**Conclusion:** Application of unsupervised machine learning to routine echocardiographic data identifies distinct heart failure phenotypes in hypertensive patients. These phenotypes demonstrate significant structural heterogeneity beyond LVEF-based classification, supporting the utility of data-driven approaches for refined cardiac phenotyping. This pilot study provides a foundation for larger prospective studies.

## INTRODUCTION

Heart failure remains a major cause of cardiovascular morbidity and mortality worldwide and is particularly prevalent among patients with longstanding systemic hypertension.^[1,2]^ Chronic pressure overload induces progressive structural and functional myocardial remodeling, resulting in heterogeneous cardiac phenotypes that range from concentric hypertrophy with preserved systolic function to eccentric remodeling with overt systolic impairment.^[3,4]^

Conventional classification of heart failure is largely based on LVEF, categorizing patients into HFrEF, HFmrEF, HFpEF and heart failure with improved ejection fraction (HFimpEF).^[1]^ Although clinically useful, this framework incompletely captures the multidimensional nature of hypertensive cardiac remodeling. Patients with similar LVEF may exhibit markedly different ventricular geometry, myocardial mass, filling dynamics, and atrial remodeling, suggesting substantial biological heterogeneity beyond systolic performance alone.[^5, 6]^

Recent advances in machine learning have enabled data-driven phenotyping of complex cardiovascular syndromes through unsupervised clustering approaches.^[7]^ By simultaneously integrating multiple echocardiographic variables, these techniques can identify latent structural and functional phenotypes not apparent using conventional classification systems.^[7]^ Such approaches have shown promise in refining heart failure taxonomy and improving mechanistic understanding of disease progression.

However, most existing studies have relied on highly curated datasets from resource-rich settings, often incorporating advanced imaging or biomarker panels that are not routinely available in many clinical environments.^[8-11]^ Limited evidence exists regarding the feasibility of applying unsupervised machine learning to routine transthoracic echocardiographic datasets, particularly in resource-limited settings.

This pilot study therefore explored whether multivariate clustering could identify clinically meaningful structural phenotypes beyond conventional LVEF-based classification.

## METHODS AND MATERIALS

### Study Design and Setting

This retrospective cross-sectional pilot study was conducted using routine transthoracic echocardiographic reports obtained from Federal Medical Centre (FMC) Umuahia, Abia State, Nigeria. The study was designed as a feasibility assessment for a larger planned dissertation investigating echocardiographic phenotyping of hypertensive heart failure using unsupervised machine learning.

### Study Population

Adult patients (≥18 years) with documented systemic hypertension and clinically established heart failure based on Framingham criteria who underwent transthoracic echocardiography between September 2025 and January 2026 were eligible.

Patients were excluded if reports were incomplete, lacked key structural or functional variables required for clustering, or represented duplicate records.

### Feature Selection and Standardization

Eleven clinically relevant echocardiographic variables reflecting ventricular geometry, systolic performance, and diastolic function were selected for clustering. All variables were standardized using z-score normalization.

### Clustering Analysis

Unsupervised clustering was performed using K-means. The optimal number of clusters was determined using elbow analysis and silhouette scoring. Principal component analysis was used for two-dimensional visualization of cluster separation.

### Statistical Analysis

Continuous variables are presented as mean ± standard deviation. Differences across clusters were assessed using the Kruskal–Wallis test. Associations between machine learning–derived clusters and conventional LVEF-based heart failure categories (HFrEF <40%, HFmrEF 40– 49%, HFpEF ≥50%) were assessed by cross-tabulation. Statistical significance was defined as p < 0.05. Analyses were performed using Python 3.x with NumPy, pandas, SciPy, and scikit-learn.

### Ethical Considerations

All data were anonymized prior to analysis. The study involved secondary analysis of retrospective clinical data and posed minimal risk to participants. Ethical approval was obtained from the ethical review committee of the FMC Umuahia.

## RESULTS

A total of 124 echocardiographic reports were initially extracted from semi-structured clinical documents. Following data preprocessing, 102 complete cases were retained for final analysis. The mean age of participants was 60.6 ± 16.8 years. Mean body mass index was 27.8 ± 6.0 kg/m^2^, and mean ejection fraction was 52.8 ± 15.1%, reflecting a heterogeneous heart failure population. Core echocardiographic variables demonstrated acceptable completeness, while right ventricular parameters were excluded because of substantial missingness (>50%).

**Table 1:** Baseline clinical and echocardiographic characteristics of the study population (n = 102)

| Variable | Mean ± SD / n (%) |
| --- | --- |
| Age (years) | 60.6 ± 16.8 |
| Weight (kg) | 73.8 ± 15.7 |
| Height (cm) | 163.3 ± 9.5 |
| BMI (kg/m<sup>2</sup>) | 27.8 ± 6.0 |
| BSA (m<sup>2</sup>) | 1.82 ± 0.21 |
| Heart rate (bpm) | 85.5 ± 15.1 |
| Systolic BP (mmHg) | 118.3 ± 24.6 |
| Diastolic BP (mmHg) | 74.2 ± 14.6 |
| LVIDd (cm) | 5.32 ± 1.31 |
| LVIDs (cm) | 3.64 ± 1.52 |
| IVSd (cm) | 1.10 ± 0.27 |
| LVPWd (cm) | 1.17 ± 0.28 |
| LVMI (g/m<sup>2</sup>) | 138.7 ± 57.8 |
| Relative wall thickness | 0.47 ± 0.17 |
| Ejection fraction (%) | 52.8 ± 15.1 |
| Fractional shortening (%) | 28.3 ± 8.0 |
| Left atrial diameter (cm) | 4.33 ± 1.00 |
| E/A ratio | 1.66 ± 1.08 |
| Mitral deceleration time (ms) | 163.2 ± 62.4 |

K-means clustering identified three distinct echocardiographic phenotypes. Optimal cluster number was supported by both elbow plot analysis and the highest silhouette score at k = 3 (0.276). The modest silhouette value likely reflects overlap within biologically continuous cardiovascular phenotypes.

**Figure 1:**
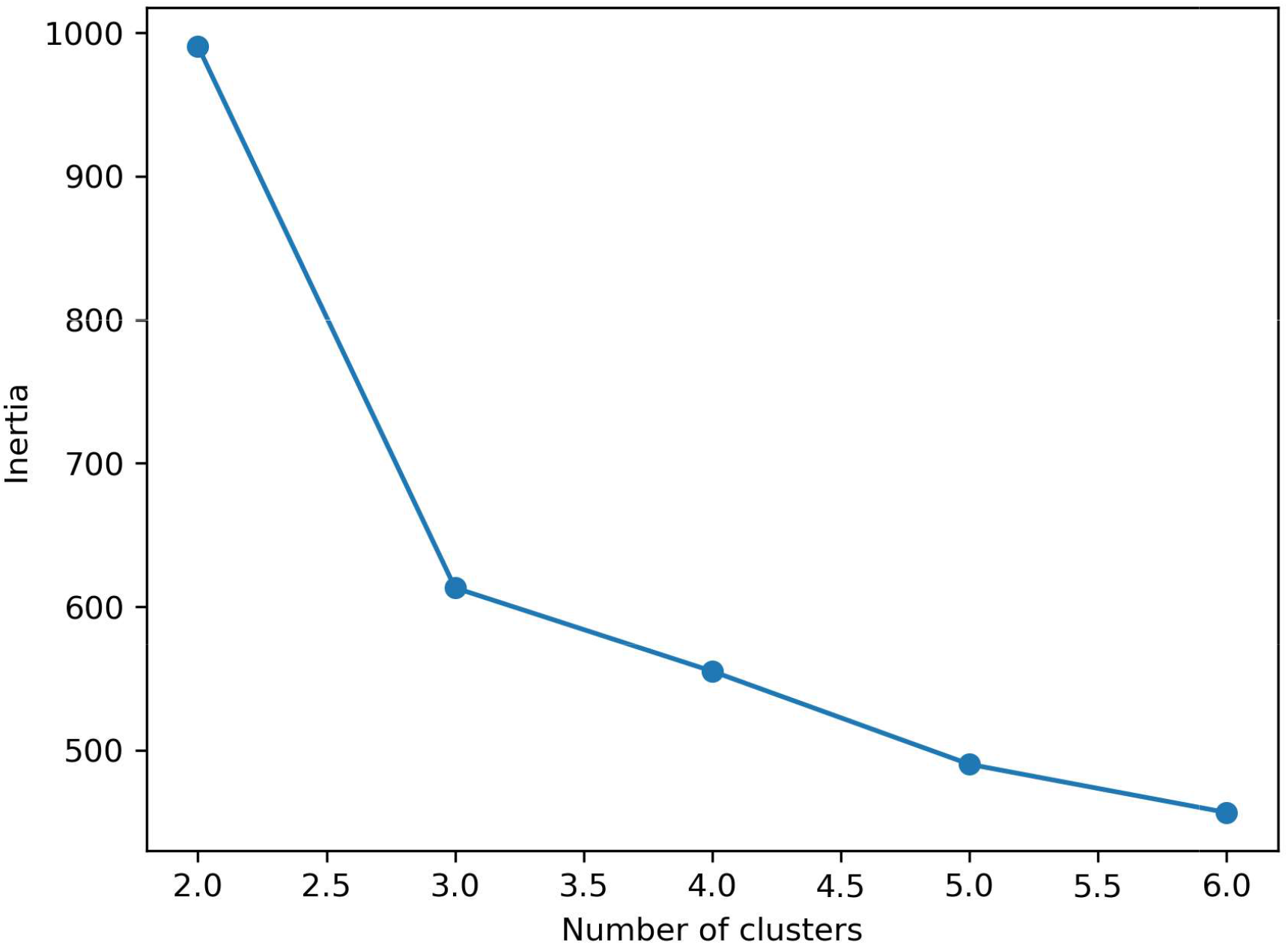
Elbow plot showing distinct inflection at k = 3

Accordingly, three distinct echocardiographic phenotypes were identified:

Cluster 0: n = 50 (49.0%)

Cluster 1: n = 37 (36.3%)

Cluster 2: n = 15 (14.7%)

Cluster 0 (Concentric HFpEF phenotype) demonstrated features consistent with concentric remodeling and HFpEF. It was marked by preserved left ventricular systolic function (EF: **∼**76%), small to normal left ventricular (LV) dimensions (LVIDd: **∼**4.45 cm), increased relative wall thickness (RWT: **∼**0.54), mild left atrial enlargement, and mild diastolic dysfunction (E/A ∼1.08)

Cluster 1 (Dilated HFrEF phenotype) demonstrated characteristics consistent with eccentric remodeling and HFrEF. It demonstrated markedly reduced LVEF (**∼**41%), significant LV dilation (LVIDd: **∼**6.6 cm, LVIDs: **∼**5.2 cm), reduced fractional shortening, lower relative wall thickness (RWT: **∼**0.31), elevated E/A ratio suggestive of restrictive filling pattern, and increased left atrial size.

Cluster 2 represented a hypertrophic phenotype with mid-range ejection fraction (Hypertrophic HFmrEF phenotype). It was characterized by intermediate LVEF (∼53%), marked concentric hypertrophy (IVSd: **∼**1.54 cm, LVPWd: **∼**1.56 cm), highest LV mass index among clusters (**∼**190 g/m^2^), increased relative wall thickness (**∼**0.64), and intermediate diastolic dysfunction parameters.

**Table 2:** Echocardiographic characteristics of identified clusters.

| **Variable** | **Cluster 0 (n = 50)** | **Cluster 1 (n = 37)** | **Cluster 2 (n = 15)** |
| --- | --- | --- | --- |
| LVIDd (cm) | 4.45 | 6.60 | 5.09 |
| LVIDs (cm) | 2.44 | 5.24 | 3.69 |
| IVSd (cm) | 1.06 | 0.99 | 1.54 |
| LVPWd (cm) | 1.16 | 1.02 | 1.56 |
| Left atrium (cm) | 3.84 | 4.74 | 4.33 |
| Ejection fraction (%) | 75.99 | 40.72 | 52.82 |
| Fractional shortening (%) | 46.72 | 21.60 | 28.27 |
| E/A ratio | 1.08 | 2.42 | 1.70 |
| Mitral deceleration time (ms) | 187.33 | 135.24 | 152.00 |
| LV mass index (g/m<sup>2</sup>) | 100.58 | 169.32 | 190.23 |
| Relative wall thickness | 0.54 | 0.31 | 0.64 |

There was partial but incomplete concordance between echocardiographic clusters and conventional LVEF-based heart failure categories. Cluster 0 showed strong alignment with HFpEF (96% HFpEF classification). Cluster 1 contained a mixed distribution, including HFrEF (49%), HFmrEF (30%), and HFpEF (21%). Cluster 2 demonstrated heterogeneous distribution across all LVEF categories.

This indicates that LVEF-based classification does not fully capture the structural heterogeneity identified through multivariate echocardiographic clustering.

Heatmap visualization demonstrated distinct patterns of left ventricular geometry, systolic function, and diastolic indices across the three clusters.

**Figure 2:**
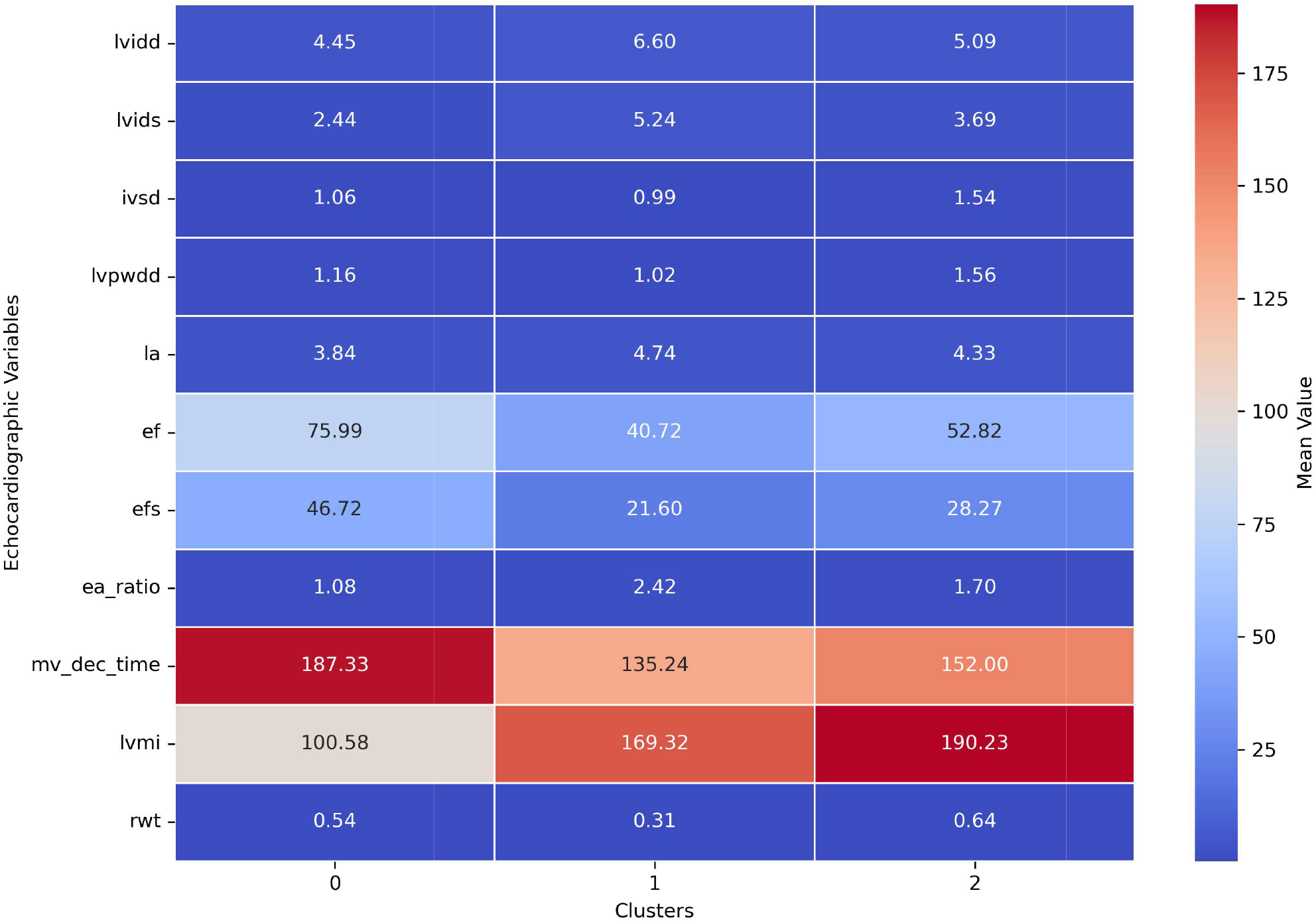
Heatmap visualization demonstrated distinct patterns of left ventricular geometry, systolic and diastolic indices across the three clusters.

All echocardiographic variables demonstrated statistically significant differences across clusters using the Kruskal–Wallis test (p < 0.001). This confirms that the identified clusters represent distinct and statistically separable echocardiographic phenotypes.

Principal component analysis showed partial but distinct separation of the three echocardiographic clusters in two-dimensional space, indicating underlying structural heterogeneity in multivariate cardiac geometry and function.

**Figure 3:**
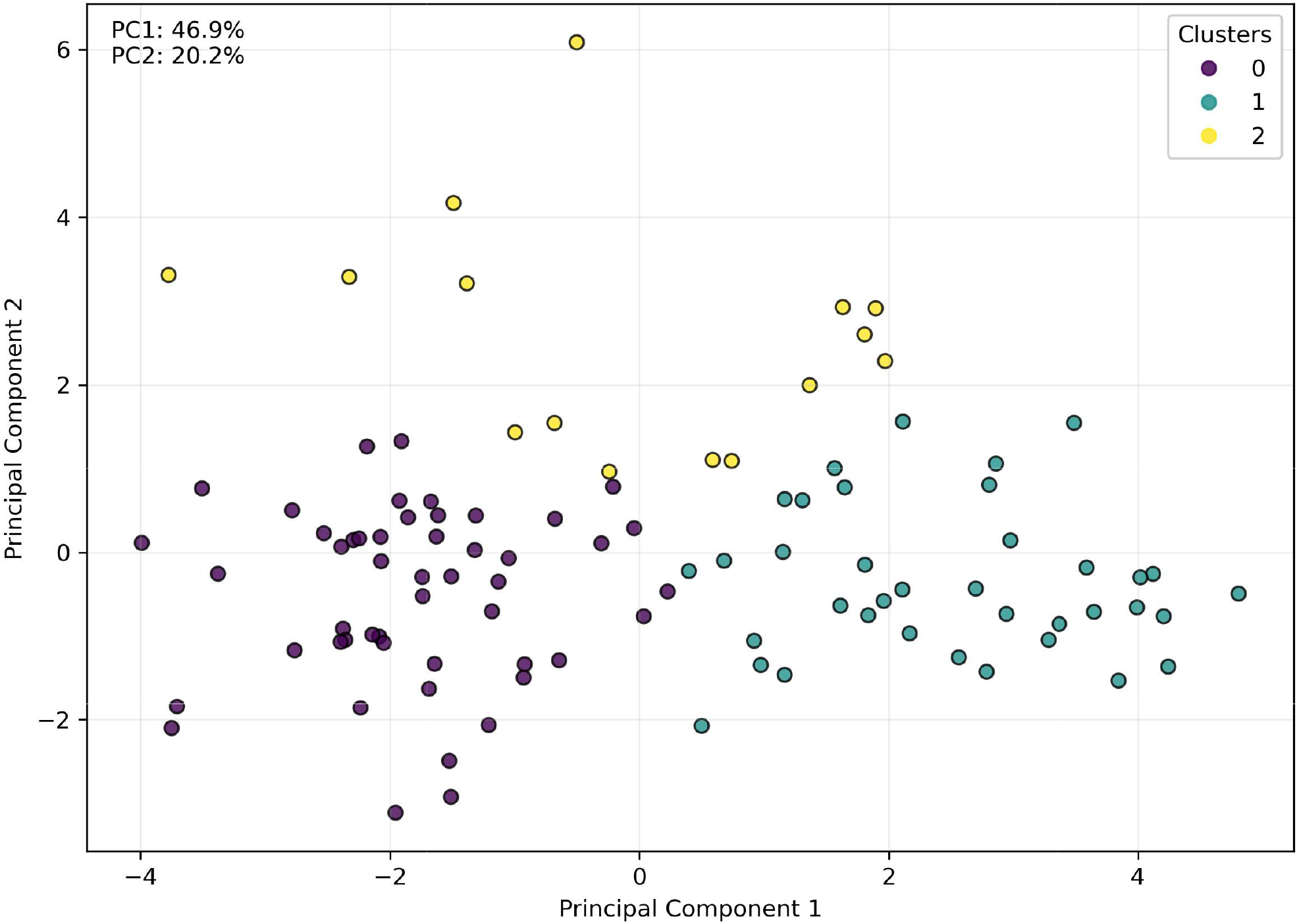
Principal component analysis showing distinct cluster separation in 2-dimensional space.

## DISCUSSION

This pilot study demonstrates the feasibility of applying unsupervised machine learning to routinely acquired echocardiographic data for phenotyping hypertensive heart failure. Three clinically interpretable phenotypes were identified: a concentric HFpEF phenotype, a dilated HFrEF phenotype, and a hypertrophic HFmrEF-like phenotype. These findings highlight substantial structural heterogeneity beyond conventional LVEF-based classification.

Although LVEF remains central to heart failure categorization, our results demonstrate incomplete concordance between LVEF-based groups and multivariate structural phenotypes, reinforcing the limitations of relying solely on systolic function for disease characterization.

Cluster 0 (Concentric HFpEF phenotype) was characterized by preserved systolic function, increased relative wall thickness, and mild left atrial enlargement. These findings are consistent with concentric remodeling secondary to long-standing hypertension.^[3,4]^ The elevated LVEF with impaired diastolic indices suggests a phenotype consistent with HFpEF. This phenotype aligns with established models of hypertensive cardiac remodeling, where pressure overload leads to concentric hypertrophy, increased myocardial stiffness, and progressive diastolic dysfunction.^[3,4]^

Cluster 1 (Dilated HFrEF phenotype) demonstrated marked left ventricular dilation, reduced systolic function, and increased left atrial size. The low relative wall thickness and reduced LVEF are consistent with eccentric remodeling and advanced systolic heart failure.^[1-5]^ The presence of elevated E/A ratios suggests restrictive filling physiology in a subset of patients, indicating more advanced diastolic impairment. This phenotype likely represents decompensated hypertensive heart disease with progression to overt systolic failure.^[1,2,5]^

Cluster 2 (Hypertrophic HFmrEF-like phenotype) exhibited marked concentric hypertrophy, elevated left ventricular mass index, and intermediate systolic function. Despite relatively preserved LVEF, structural indices indicate advanced myocardial remodeling. This phenotype likely represents a transitional stage between HFpEF and HFrEF, consistent with progressive hypertensive cardiomyopathy.^[1,2,5,12]^ The coexistence of increased wall thickness and intermediate LVEF suggests a high-risk subgroup with mixed functional impairment.

A major finding of this study is the incomplete overlap between unsupervised echocardiographic clusters and conventional LVEF-based heart failure classification.

While Cluster 0 showed strong alignment with HFpEF (96%), Clusters 1 and 2 demonstrated substantial heterogeneity across heart failure categories. Notably, patients classified as HFpEF or HFmrEF by LVEF criteria were distributed across multiple structural phenotypes.

This discordance reinforces a growing body of evidence that LVEF-based classification alone does not adequately reflect underlying myocardial remodeling patterns. Instead, heart failure in hypertensive patients appears to exist along a multidimensional spectrum defined by geometry, mass, and diastolic function rather than systolic function alone.

All selected echocardiographic variables demonstrated statistically significant differences across clusters (p < 0.001), confirming that the identified phenotypes are not random artifacts of clustering but represent underlying structural distinctions.

The combination of the elbow method and silhouette analysis supported an optimal cluster solution of three although the moderate silhouette score (0.276) reflects expected overlap in biological systems rather than methodological weakness. In cardiovascular phenotyping, moderate cluster separation is common due to continuous rather than categorical disease progression.^[6,7]^

Previous studies on heart failure phenotyping have largely focused on clinical or biomarker-based clustering approaches.^[6,7,8]^ Fewer studies have utilized high-dimensional echocardiographic structural data, particularly in hypertensive populations.

The present study extends prior work by demonstrating that routine echocardiographic parameters alone are sufficient to generate clinically meaningful phenotypes using unsupervised learning. This is particularly relevant in resource-limited settings where advanced imaging modalities may not be readily available.

### Clinical implications

The findings of this pilot study have several important implications:

- EF alone is insufficient to characterize the full spectrum of hypertensive heart failure phenotypes.
- Routine echocardiographic data can be repurposed for advanced phenotyping using machine learning techniques.
- Structural phenotypes may offer additional prognostic value beyond traditional LVEF categories.
- The identified clusters provide a foundation for prospective validation and outcome-based studies.

### Limitations

This pilot study has several limitations. First, the sample size is modest, and findings require validation in larger cohorts. Second, right ventricular functional parameters were incompletely captured due to missing data, limiting comprehensive biventricular phenotyping. Third, the retrospective nature of data extraction introduces potential variability in measurement quality. Finally, external validation across multiple centers was not performed.

## Conclusion

This study demonstrates the feasibility and clinical utility of unsupervised machine learning applied to routine echocardiographic data in hypertensive heart failure. Three reproducible and clinically meaningful phenotypes were identified, revealing significant heterogeneity within LVEF-based heart failure classification. These findings support a shift toward multidimensional echocardiographic phenotyping as a complementary approach to conventional classification systems and provide a strong foundation for future large-scale prospective studies.

## Data Availability

All data produced in the present study are available upon reasonable request to the authors.

